# Derivation and validation of a prognostic model for predicting in-hospital mortality in patients admitted with COVID-19 in Wuhan, China: the PLANS (Platelet Lymphocyte Age Neutrophil Sex) model

**DOI:** 10.1101/2020.05.13.20100370

**Authors:** Jiong Li, Yuntao Chen, Shujing Chen, Sihua Wang, Dingyu Zhang, Junfeng Wang, Douwe Postmus, Hesong Zeng, Guoyou Qin, Yin Shen, Jinjun Jiang, Yongfu Yu

**Affiliations:** MOE-Shanghai Key Laboratory of Children’s Environmental Health, Xin Hua Hospital Affiliated to Shanghai Jiao Tong University School of Medicine, Shanghai, China; Department of Epidemiology, University Medical Center Groningen, Groningen, The Netherlands; Department of Pulmonary and Critical Care Medicine, Zhongshan Hospital, Fudan University, Shanghai, China; Department of Thoracic Surgery, Union Hospital, Tongji Medical College, Huazhong University of Science and Technology, Wuhan, China; Department of Tuberculosis and Respiratory Disease, Jinyintan Hospital, Wuhan, China; Julius Center for Health Science and Primary Care, University Medical Center Utrecht, Utrecht University, Utrecht, The Netherlands; Department of Cardiology, Tongji Hospital, School of Medicine, Huazhong University of Science and Technology, Wuhan, China; Department of Biostatistics, School of Public Health, and The Key Laboratory of Public Health Safety of Ministry of Education, Fudan University, Shanghai, China; Eye Center, Medical Research Institute, Wuhan University Renmin Hospital, Wuhan University, Wuhan, China

**Keywords:** COVID-19, in-hospital mortality, prognostic model

## Abstract

**OBJECTIVE:** To develop and validate a prognostic model for in-hospital mortality in COVID-19 patients using routinely collected demographic and clinical characteristics.

**DESIGN:** Multicenter, retrospective cohort study.

**SETTING:** Jinyintan Hospital, Union Hospital, and Tongji Hosptial in Wuhan, China.

**PARTICIPANTS:** A pooled derivation cohort of 1008 COVID-19 patients from Jinyintan Hospital, Union Hospital in Wuhan and an external validation cohort of 1031 patients from Tongji Hospital in Wuhan, China.

**MAIN OUTCOME MEASURES:** Outcome of interest was in-hospital mortality, treating discharged alive from hospital as the competing event. Fine-Gray models, using backward elimination for inclusion of predictor variables and allowing non-linear effects of continuous variables, were used to derive a prognostic model for predicting in-hospital mortality among COVID-19 patients. Internal validation was implemented to check model overfitting using bootstrap approach. External validation to a separate hospital was implemented to evaluate the generalizability of the model.

**RESULTS:** The derivation cohort was a case-mix of mild-to-severe hospitalized COVID-19 patients (n=1008, 43.6% females, median age 55). The final model (PLANS), including five predictor variables of platelet count, lymphocyte count, age, neutrophil count, and sex, had an excellent predictive performance (optimism-adjusted C-index: 0.85, 95% CI: 0.83 to 0.87; averaged calibration slope: 0.95, 95% CI: 0.82 to 1.08). Internal validation showed little overfitting. External validation using an independent cohort (n=1031, 47.8% female, median age 63) demonstrated excellent predictive performance (C-index: 0.87, 95% CI: 0.85 to 0.89; calibration slope: 1.02, 95% CI: 0.92 to 1.12). The averaged predicted survival curves were close to the observed survival curves across patients with different risk profiles.

**CONCLUSIONS:** The PLANS model based on the five routinely collected demographic and clinical characteristics (platelet count, lymphocyte count, age, neutrophil count, and sex) showed excellent discriminative and calibration accuracy in predicting in-hospital mortality in COVID-19 patients. This prognostic model would assist clinicians in better triaging patients and allocating healthcare resources to reduce COVID-19 fatality.

## INTRODUCTION

The novel coronavirus disease (COVID-19) has become a pandemic worldwide since its first outbreak in Wuhan, China since December 2019.^1^ As of May 12, 2020, more than 4 million cases are confirmed in more than 200 countries, including 283 271 deaths.^2^ Due to the high contagiousness and rapid progression of the disease, healthcare demand, in particular for critical care capacities, has often been overwhelming even in high-income areas.^3^ Good support tools are needed for clinicians and other healthcare workers to respond promptly to urgent situations. It is crucial to accurately select severe patients for targeted treatment. For example, while it is essential to increase the intensive care unit (ICU) capacities and staff, ICU triage may be critical to prioritize severe patients for intensive care.^4^ Therefore, early stratification of patients will facilitate targeted supportive care and appropriate allocation of medical resources.

Prognostic model that combines several clinical or non-clinical variables to estimate the future health outcomes of an individual could be a useful tool^5^ To respond quickly to the health crisis of COVID-19, a prognostic model based on robust evidence could be used as a simple and inexpensive tool to assist doctors in triaging the patients in the first place, which in turn may mitigate the burden of overwhelmed healthcare system and better allocate limited healthcare resources to reduce COVID-19 fatality.^6^ Currently, several clinical prognostic models have been developed for COVID-19 patients.^7 8^ However, the quality of these models has been criticized and was prone to bias due to unrepresentativeness of patient population, lack of external validation, inappropriate statistical analyses, or poor reporting.^7^ Two of these prognostic models have been constructed with promising predictive performance for predicting mortality.^9 10^ However, they may not be highly reliable due to relatively small derivation cohorts (189 to 296 patients) and external validation cohorts (19 to 165 patients).In addition, some of the predictor variables included in the final models may not be routinely measured, which in turn limited the implementation of the model to clinical practice. Furthermore, the model generalizability to different settings is rarely considered in the model derivation. Given the fact of the ongoing worldwide pandemic, a reliable prognostic model should not only be applied in the local setting but also can be generalized in different settings after updating.

In this study, we aimed to develop a prognostic model to predict in-hospital mortality in COVID–19 patients using routinely measured demographic and clinical characteristics from two state–designated hospitals for COVID–19 treatment in Wuhan, China. We also validated this model in another independent hospital in Wuhan. Furthermore, we developed two updated prognostic models accounting for different characteristics in patients from New York in the USA and Lombardy in Italy.

## METHODS

### Study cohorts

#### Derivation cohort

The derivation cohort included 1008 COVID–19 patients admitted at Jinyintan Hospital (n=763) and Union Hospital (n=245) in Wuhan, China from January 1 to February 10, 2020 and all patients were followed up to March 20, 2020. The Jinyintan hospital had mostly severe patients while Union Hospital had mostly mild patients, thus the cohort consisted of a case-mix of mild-severe COVID-19 patients.

#### Validation cohort

The validation cohort included 1031 COVID–19 patients aged ≥ 18 years at Tongji Hospital in Wuhan, China from January 14 to March 8, 2020. Since this cohort was designed to assess the potential risk factors related to acute cardiac injury in COVID–19 patients, the cohort did not include patients with the stage of chronic kidney disease ≥ 4, chronic heart failure in the decompensatory stage, acute myocardial infarction during hospitalization, or having missing information on hypersensitive cardiac troponin I. Patients were followed up to March 30, 2020.

### Data collection

A trained team of physicians retrospectively reviewed clinical electronic medical records and laboratory findings for all the patients. All patients met the diagnostic criteria according to the WHO interim guidance.^11^ In the derivation cohort, we collected data on age, sex, the dates of admission and discharge or death, complete blood count at admission (neutrophil, lymphocyte, platelet count, haemoglobin), current smoking status (no, yes), chronic disease history (hypertension, digestive disease, kidney disease, coronary heart disease(CHD), chronic pulmonary disease, cerebrovascular disease, diabetes, thyroid disease, malignancy, and other diseases). In the validation cohort, we collected data on age, sex, the dates of admission and discharge or death, complete blood count at admission (neutrophil, lymphocyte, platelet count), chronic disease history (hypertension, diabetes, CHD). All data were reviewed and collected by two physicians and a third researcher adjudicated any difference in interpretation between the two physicians.

### Outcome and candidate predictors

The outcome of interest was in-hospital mortality. Length of hospital stay (LOS) was defined as the time from hospital admission to either discharged alive or death. The follow-up started from hospital admission and ended at death, discharged alive, or 30–day after hospital admission (administrative censoring at 30-day after hospital admission), whichever came first. Candidate predictor variables were selected according to clinical knowledge, literature,^7 12^ and data availability, including age, sex, neutrophil count, lymphocyte count, platelet count, current smoke status, comorbidities (hypertension, CHD, diabetes, cerebrovascular disease, and malignancy). While current smoking status was not considered due to high proportion of missing data in the derivation cohort (46.3% missing), information on all other candidate predictor variables and outcome was complete for data analysis.

### Model derivation

Fine-Gray models were used to develop the prognostic model, treating discharged alive from hospital as a competing event.^13^ The prognostic model derivation consisted of a prognostic index (PI) that captured the effect of the predictor variables on cumulative mortality rate, and a baseline survival that determined the survival of an “average” patient, i.e., a patient with the average value of PI.

First, uni-variable Fine-Gray models with fractional polynomials (maximum permissible degree 1) were performed to investigate the functional form for the continuous variables. Second, a multivariable Fine-Gray model with all the predictors was built. Backward elimination was applied to do the variable selection with significant level setting to 0.05, resulting in a final model in this step. PI was then calculated based on the combination of β coefficients and values of the corresponding predictors.

The baseline survival S_0_(t) corresponds to the survival of an “average” patient with the average value of PI. The survival of other patients can be computed via the formula:
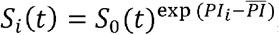 where *PI_i_* is the 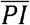 of patient i and PIis the average value of PI in the derivation cohort.

Details about the inplementation and estimates of the Fine-Gray model, see the Supplementary Appendix S1.

### Model performance and internal validation

Model performance was assessed in terms of discrimination and calibration. Discrimination was assessed using the concordance statistic (C–index).^14^ Calibration was assessed jointly by calibration slope and calibration plot. Calibration slope is a measure to estimate the regression coefficient on the PI in the validation dataset (equaling to exactly 1 in the derivation data).^15^ In the calibration plot, the averaged predicted survival curves estimated by the proposed prognostic model were compared with the averaged observed survival curves across several risk groups. The risk group was based on patients’ PI (thresholds: 15^th^,50^th^ and 84^th^ percentiles).^16^

We performed internal validation to estimate the optimism (the level of model overfitting) and adjusted measures of C–index and calibration slope by bootstrapping 1000 samples of the original data. Details about the implementation of bootstrap can be found in Supplementary Appendix S2. Average calibration slope in the internal validation was obtained to be a uniform shrinkage factor. We multiplied the shrinkage factor by the raw PI (PI in the model derivation step) to obtain optimism-adjusted PI. Lastly, we developed the final model by re-estimating the baseline survival probabilities based on the optimism-adjusted PI.

### External validation

The final model was applied to each patient in the external validation cohort. PI was then calculated based on the combination of β coefficients and the corresponding predictor values of every patient in the validation cohort. The discriminative accuracy of the proposed model was evaluated using C–index and visually checked by the distribution of PIs. The calibration accuracy of the proposed model was assessed using calibration slope and visually checked by plotting agreement between predicted and observed survival curves across four risk groups as done in the derivation cohort.

### Model update

The proposed model may not be directly applied to other areas where the distribution of predictive factors may be different from that in Wuhan. For instance, New York of USA and Lombardy of Italy could have a different distribution of preditor variables compared with Wuhan. Therefore, we used entropy balancing to update proposed model to generalize to their settings.^17^ First, entropy balancing approach was implemented to estimate a weight that made our derivation data comparable with the New York cohort in terms of the distribution of age, sex, hypertension, CHD, diabetes and malignancy. Second, a weighted Cox regression was used estimate the baseline survival of the “average” patient in New York by offsetting the PI. Last, the updated prognostic model for New York can be obtained via 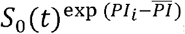 where S_0_(t) is a New York specific baseline survival, *PI_i_* is the PI of patient i and 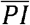 is the PI of the average patient in New York, which was assumed the same as that in Wuhan. Same procedures were implemented to obtain the updated model in Lombardy.

### Statements about reporting and evaluation of our prognostic model

The reporting of this prognostic model study followed Transparent Reporting of a multivariable prediction model for Individual Prognosis Or Diagnosis (TRIPOD) statement (Supplementary).^18^ The risk of bias of the prognostic model was independently assessed by an expert (JW, who did not take part in the model development and validation) using PROBAST (prediction model risk of bias assessment tool).^19^

### Patient and public involvement

No patients were involved in setting the research question or the outcome measures, nor were they involved in developing plans for design or implementation of the study. No patients were asked to advise on interpretation or writing up of results. There are no plans to disseminate the results of the research to study patients or the relevant patient community.

## RESULTS

### Patient population

Summary statistics of the patient characteristics at hospital admission are provided in Table 1. In the derivation cohort, the median age of 1008 patients was 55 (interquartile range [IQR] 44-65, youngest at 14 years of age and oldest at 98 years) and 43.6% patients were females.During a median LOS of 12 days (IQR 8-16), 211 (20.9%) died and the youngest and oldest deceased patients were 14 and 98 years old, respectively. There were 438 (43.5%) patients with one or more comorbidities. Hypertension (N=232, 23.0%), diabetes (N=110, 10.9%), chronic digestive disease (N=78, 7.7%), and chronic pulmonary disease (N=40, 4.0%) were among the most frequent comorbidities.

In the validation cohort, the 1031 patients included were older (63, IQR 52-70), had more females (47.8%), and were more prevalent with hypertension (N=383, 37.1%), CHD (N=83, 8.1%) and diabetes (N=189, 18.3%), compared to the derivation cohort. Patients had a longer LOS (19, IQR 11-27). The survival probability of patients in the validation cohort was slightly higher compared with the derivation cohort (Figure S1).

**Table 1.** Basic characteristics

|  | Derivation cohort (n=1008) | Validation cohort (n=1031) |
| --- | --- | --- |
| Age, years | 55 (44-65) | 63 (52-70) |
| Sex, female | 439 (43.6%) | 493 (47.8%) |
| Current smoke status<sup>a</sup> | 57 (10.5%) | - |
| Neutrophil count, $\times 10^9/L$ | 4.40 (2.79-6.96) | 3.90 (2.78-5.68) |
| Lymphocyte count<sup>b</sup>, $\times 10^9/L$ | 0.95 (0.61-1.34) | 1.07 (0.70-1.49) |
| Platelet count<sup>c</sup>, $\times 10^9/L$ | 194 (145-256) | 219 (164-288) |
| Haemoglobin<sup>d</sup>, g/L | 126 (115-138) | - |
| Chronic pulmonary disease | 40 (4.0%) | - |
| Hypertension | 232 (23.0%) | 383 (37.1%) |
| Coronary heart disease | 32 (3.2%) | 83 (8.1%) |
| Diabetes | 110 (10.9%) | 189 (18.3%) |
| Thyroid disease | 31 (3.1%) | - |
| Chronic digestive disease | 78 (7.7%) | - |
| Cerebrovascular disease | 22 (2.2%) | - |
| Chronic kidney disease | 25 (2.5%) | - |
| Malignancy | 31 (3.1%) | - |

<sup>a</sup> Current smoke status was missing in 467 (46.3%) patients in the derivation cohort
<sup>b</sup> Lymphocyte count was missing in 1 patient in the validation cohort
<sup>c</sup> Platelet count was missing in 2 patients in the validation cohort
<sup>d</sup> Haemoglobin was missing in 376 (37.3%) patients in the derivation cohort

### Coding of predictors

Categorical predictors (sex, hypertension, CHD, diabetes, cerebrovascular disease and malignancy) were coded as dummy variables. Among continuous predictors, we did not observe obvious violation of linearity assumption for age, neutrophil and platelet count. We observed a non-linear relation between outcome and lymphocyte count, especially for lymphocyte count < 2 × 10^9^/L. Therefore, we included the transformed lymphocyte count (square root of the lymphocyte count) in the model according to the results of fractional polynomial analyses.

### Model derivation and internal validation

The PLANS model included five predictors: platelet count, lymphocyte count, age, neutrophil count, and sex (Table 2). In-hospital mortality was associated with older age, being male, higher neutrophil, lower lymphocyte and lower platelet count. This model showed excellent apparent discriminative ability (C-index: 0.85, 95% CI: 0.83 to 0.88). After adjusting for overfitting, the model maintained excellent discriminative accuracy (optimism-adjusted C-index: 0.85, 95% CI: 0.83 to 0.87). The average calibration slope (uniform shrinkage factor) was 0.95 (95% CI: 0.82 to 1.08), again suggesting little model overfit. The final PI was calculated as 0.95 (uniform shrinkage factor) times the raw PI and the formula for final PI was structured as

PI = –0.002* Platelet – 2.399 * Lymphocyte+ 0.044 * Age + 0.127 * Neutrophil + 0.468 * Sex (Formula 1)

– Platelet: ×10^9^/L
– Lymphocyte: ×10^9^/L, transformed to lymphocyte ∧0.5
– Age: in years
– Neutrophil: se ×10^9^/L
– Sex: female=0; male=1

The distribution of final PI suggested good discriminative ability of our model (upper panel of Figure 1).The relationship between PI and 7-day, 14-day and 30-day survival probabilities are presented in Figure 2. While we observed a slight underestimate of survival in the highest risk group, the agreement between predicted survival curves and the observed survival curves in the other risk groups suggested good calibration of our model (left panel of Figure 3).The final formula for calculating survival probability for patient i is 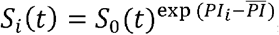 where S_0_(t) is the survival probability of the “average” patient with PI equaling to 0.5662 (Supplementary Table S1); *PI_i_* is the prognostic index of patient i and can be calculated by Formula 1; is the mean value of PIs in the derivation cohort and is 0.5662. A patient example of how the PLANS model can be applied in the real clinical practice is depicted in Box 1

#### Box 1: Final prognostic model (PLANS)

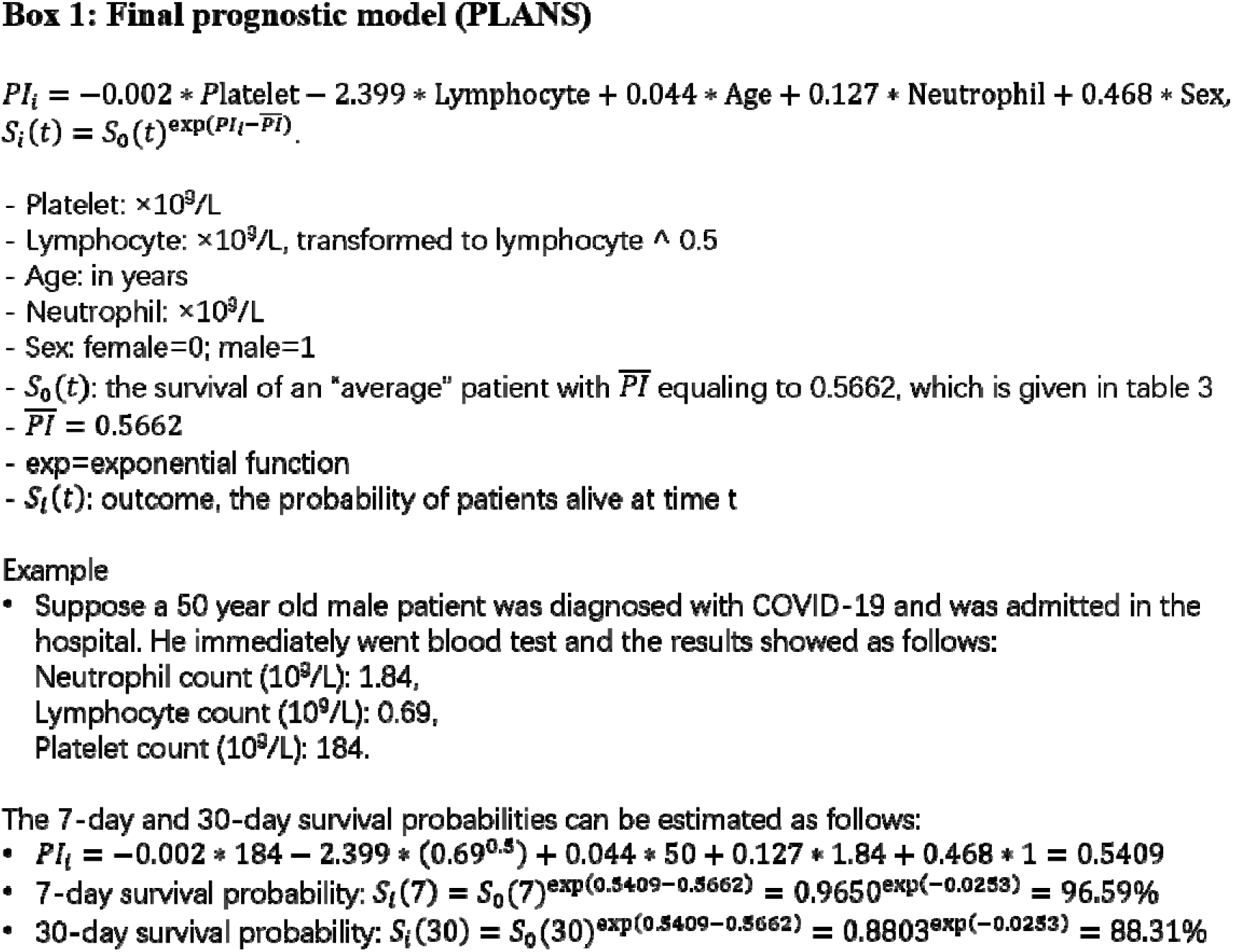

**Table 2.** Results from multi-variable Fine-Gray model

| Variables | Coding | Coefficient | 95% CI | P |
| --- | --- | --- | --- | --- |
| Age | =x<sup>†</sup> | 0.046 | 0.036 - 0.057 | <0.001 |
| Sex | Dummy (0=Female, 1=Male) | 0.490 | 0.179 - 0.802 | 0.002 |
| Neutrophil count | =x | 0.133 | 0.109 - 0.156 | <0.001 |
| Lymphocyte count | Lymphocyte count ^ 0.5 | -2.514 | -3.192- -1.835 | <0.001 |
| Platelet count | =x | -0.002 | -0.004 - -0.001 | 0.028 |

<sup>†</sup> x stands for original value.

**Figure 1.**
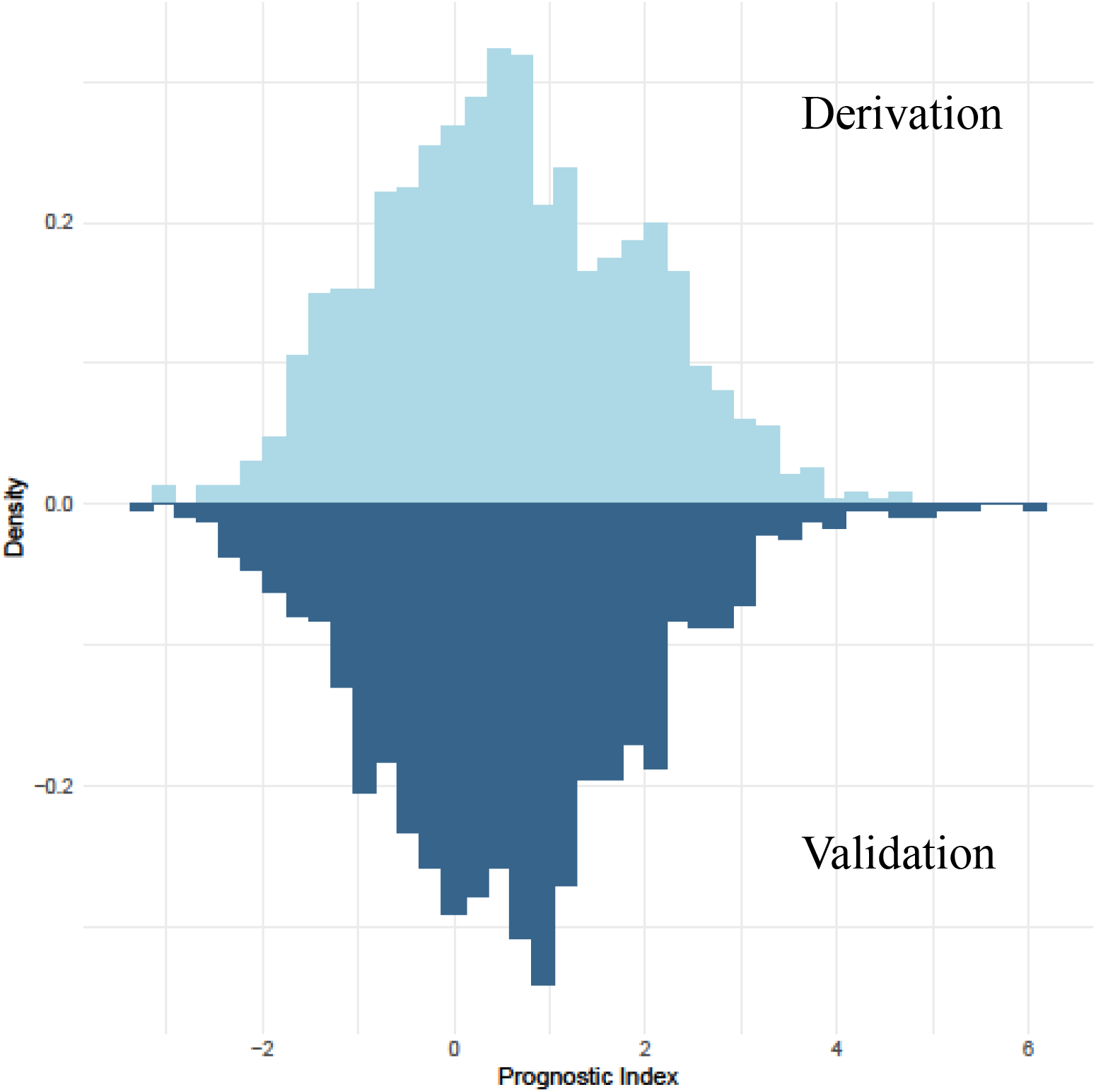
Distribution of the prognostic index of the prognostic model in the derivation and validation cohort

**Figure 2.**
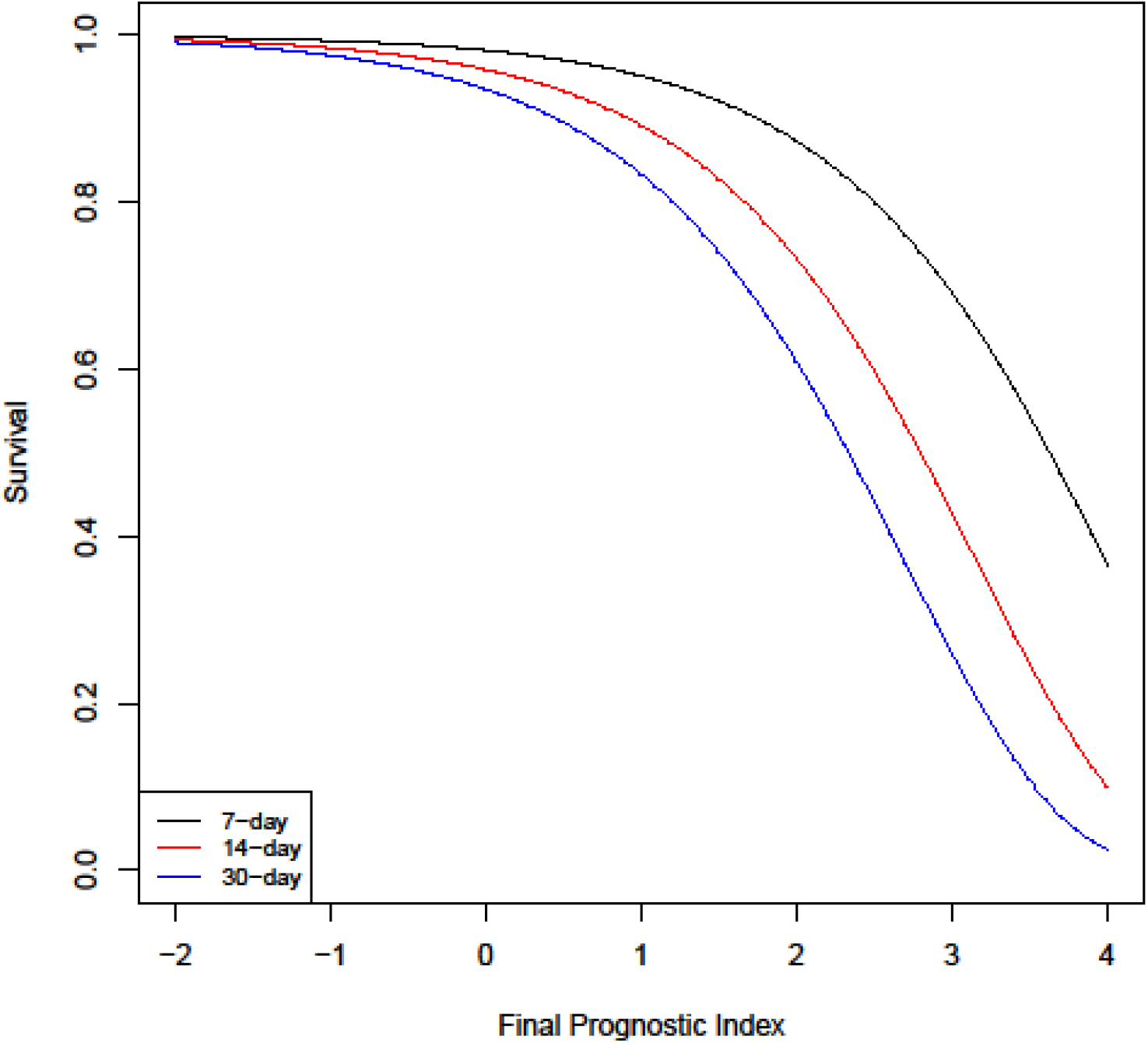
Prediction of 7-day, 14-day and 30-day survival probability versus final prognostic index

### External validation

We applied the PLANS model to the independent cohort of 1031 patients from Tongji Hospital. The distribution of the PIs in the validation cohort was very similar to that in the derivation cohort, suggesting that the excellent discriminative accuracy of our model maintained in the validation cohort (Figure 1). The resulting C-index showed excellent discriminative accuracy of our model (C-index: 0.87, 95% CI: 0.85 to 0.89). Regarding the calibration accuracy, our model slightly underestimated survival in each risk group (right panel of Figure 3). Details about the thresholds and corresponding proportion and death toll included in each risk group are provided in Supplementary Table S2. Jointly considering a close-to-one calibration slope (1.02, 95% CI: 0.92 to 1.12) and good agreement between predicted and observed survival curves, our model still suggested good calibration accuracy in the validation cohort.

**Figure 3.**
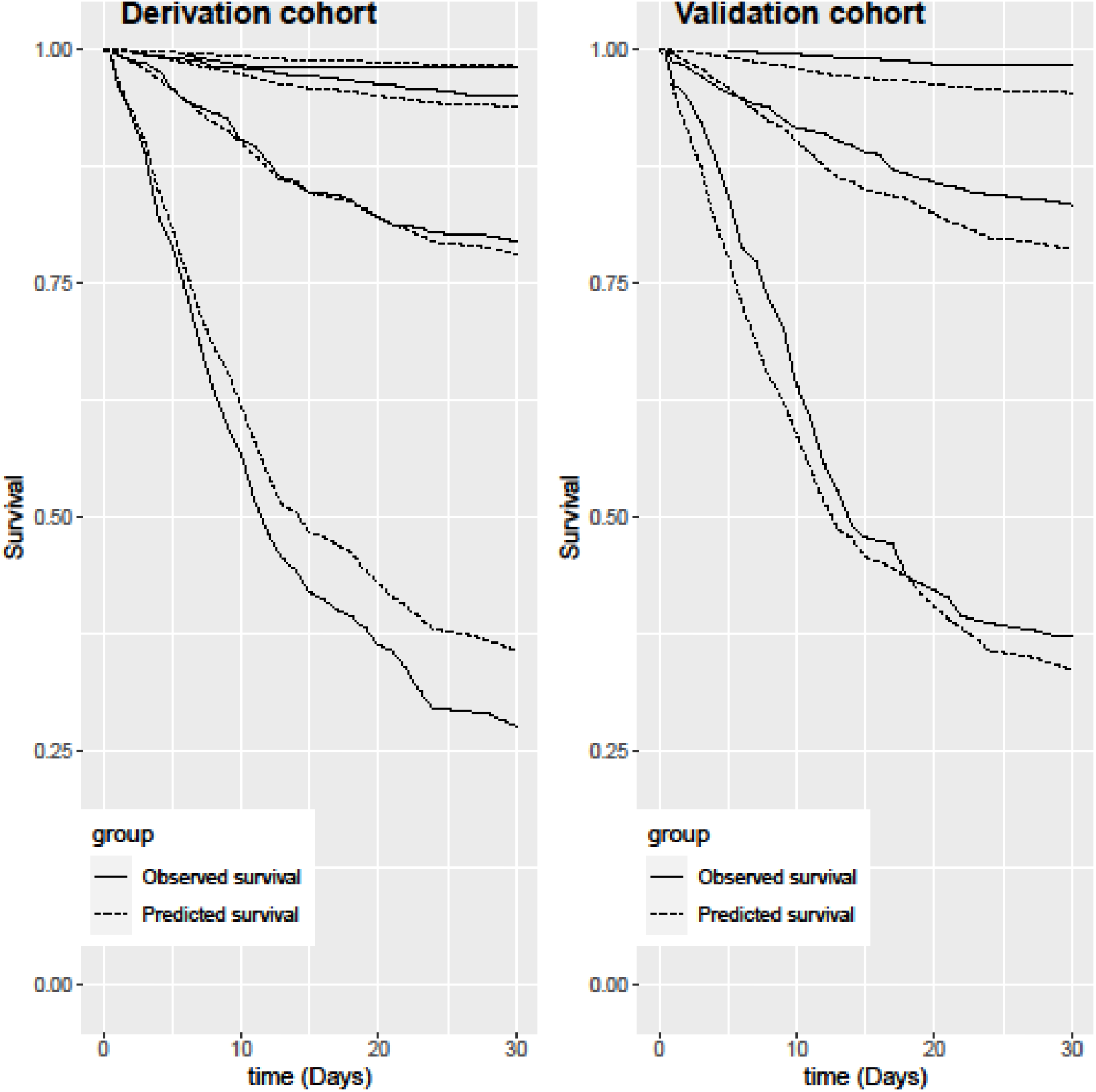
Predicted vs. Observed survival per risk group in thederivation and validation cohort

### Model update

Compared with those in Wuhan, patients in New York and Lombardy were older and prevalent with comorbidities (Supplementary Table S3). While Wuhan and New York had similar gender composition, Lombardy had a much higher of proportion of males (Supplementary Table S3). With the same PI equalling to 0.566, patients in New York had slightly better survival, while those in Lombardy had poorer survival compared with those in our derivation cohort in Wuhan. The final formula for calculating the survival probability for patient i is 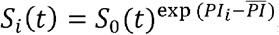 where S_(t)_ is the survival probabilities of the “average” patient in New York or Lombardy and is given in Supplementary Table S4 and Table S5; respectively; PIt is the prognostic index of patient i and can be calculated by formula 1; 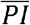 is the mean value of PIs in the derivation cohort and is 0.5662.

### Methodology quality assessment

According to the PROBAST, the proposed model was rated as low risk of bias in all four domains: 17 of the total signaling questions were “Yes” and 3 were “Probably Yes”. Rationales of answers were shown in Table S6.

## DISCUSSION

We developed a prognostic model (PLANS), using clinical readily available measures of platelet count, lymphocyte count, age, neutrophil count, and sex, to predict in-hospital mortality for COVID-19 patients in two retrospective cohorts in Wuhan, China. This model was first internally validated using bootstrap and then externally validated in an independent cohort in Wuhan. The PLANS model showed excellent discriminative and calibration accuracy. We further updated the PLANS model using summary statistics of the New York and Lombardy population, producing two adapted models for potential use in those two areas.

All the five predictors are routinely collected and some of them have been already well established as the risk factors for in-hospital mortality in previous studies.^20^ Recent studies from Italy, the USA, and China^21-23^ have also reported that advanced age was a strong predictor of in-hospital mortality as suggested in our study. Compared to previous studies,^24 25^ our study had a more balanced gender composition. Our finding that male gender was associated with increased in-hospital mortality provided further evidence to support the hypothesis of male’s vulnerability to COVID-19.^26 27^ Our study further confirmed that poor prognosis was associated with higher neutrophil and lower lymphocyte count.^28^ On top of that, lymphopenia was found to have a non-linear relation with in-hospital mortality. A meta-analysis of nine studies had reported that thrombocytopenia was significantly associated with the severity of COVID-19 disease, but heterogeneity between studies was high.^29^ Given a relatively large sample size and longer follow-up, our study indicated thrombocytopenia was associated with a higher risk of in-hospital mortality. Other studies have shown that several comorbidities (hypertension, diabetes, and coronary heart disease) were associated with poor prognosis.^21 30^ While none of the comorbidities were included in our model, we found that diabetes status would be incorporated when we excluded age from our model. It is plausible as the prevalence of most comorbidities, in particular diabetes, increases with age.^22^

Since the outbreak of COVID-19 in Wuhan, a number of prognostic models have been established.^7^ A comprehensive systematic review conducted by Wynants and colleagues found that most of these models were of high risk of bias due to several methodological limitations from participant domain to analysis domain.^7^ Compared to the previous models, the PLANS model has several strengths. Our derivation cohort had a relatively large sample size with complete information on candidate predictors. While duration of follow up was unclear in most of the previous studies, the patients in our study were followed over a relatively long period, allowing us to perform a time-to-event analysis to predict in-hospital mortality by administratively censoring at 30 days after admission to hospital. A competing risk analysis treating discharged alive as a competing event was done in this study to avoid overestimation of mortality. The similar distribution of age and sex in our study to recent large international reports^31 32^ indicates good representativeness of the patient population. External validation of the PLANS model to a large sample of patients showed excellent discrimination and calibration accuracy, indicating the generalizability of the PLANS model in the same city. Furthermore, we explored the possibility of generalizing the PLANS model to New York and Lombardy by using the published summary statistics. Though the adapted models are not recommended being applied before external validation, it might still be a good initiative to develop them and make use of them in the areas where the pandemic is still prevailing. The assessment of our model using PROBAST showed our model was of low risk of bias and out-performed currently available models.

Several limitations should be noted. First, like most of the previous datasets and two main initiatives which created protocols for the investigators, namely, the ‘International Severe Acute Respiratory and emerging Infectious Consortium (ISARIC)’ (isaric.tghn.org) and the ‘Lean European Open Survey on SARS-CoV-2 Infected Patients (LOESS)’ (leoss.net), we only include closed (discharged or dead) COVID-19 cases. However, the resulting bias of unrepresentative sample could be largely offset by the long period of follow-up time. The patients excluded from our study at least stayed in hospital for 40 days (the period from inclusion deadline to the end of follow-up). These patients may be quite heterogeneous from those included in our study. Second, we did have missing data on current smoking status for some patients. Inclusion of smoking status into the current model might improve the model performance. However, a reliable mechanism under the association between smoking and negative progression of COVID-19 is still missing.^33^ Third, some potential risk factors confirmed by previous studies, such as D-dimer,^3^ 28 were not available in our study. However, considering the practicality and validity in clinical application, a simple and interpretable model is usually preferred.^34^ In addition, our model showed promising performances with five routinely available predictors, balancing the trade-off between model performance and model practicality.

### Implication for practice

The availability of a prognostic model that can accurately predict in-hospital mortality in COVID-19 patients upon admission to hospital has important implications for practice and policy. The PLANS model may assist physicians to early stratify the patients according to the estimated mortality at 7-day (14-day or 30-day) after admission, thus giving patients targeted supporting care and better allocating the limited medical facilities (e.g. ventilators), especially when critical care capacities are overwhelmed. Several studies showed that physicians have been experiencing guilt when they make clinical decisions that contravene the morals of those making them, e.g. one ventilator, two patients.^35 36^ The PLANS model might be useful to be incorporated into a protocol to assist physicians in making those difficult decisions. Our findings from the model update suggest that our model might be generalized to different countries as well. The model could be validated in the first place and then be used directly if it performs well or after some update according to local settings.^37^

### Conclusion and future research

The PLANS model can be a guidance model for Chinese hospitals in case of the resurgence of COVID-19. It can also be a useful tool for predicting mortality or triage patients in the countries where COVID-19 is still a pandemic after being validated in their settings. Future studies are warranted about the impact of the PLANS model on clinical practice and decision.

## Data Availability

Data sharing: No additional data available.

### WHAT IS ALREADY KNOWN ON THIS TOPIC

The global pandemic of coronavirus disease 2019 (COVID-19) is still under rapid progression worldwide and causes thousands of deaths daily.

Previous published prognostic models have been criticized and are prone to bias due to unrepresentativeness of patient population, lack of external validation, inappropriate statistical analyses, or poor reporting.

A high-quality and easy-to-use prognostic model to predict in-hospital mortality for COVID-19 patients could support physicians to make better clinical desicions.

### WHAT THIS STUDY ADDS

Using a pooled derivation cohort of 1008 COVID-19 patients from Jinyintan Hospital, Union Hospital in Wuhan and an external validation cohort of 1031 patients from Tongji Hospital in Wuhan, China, we developed a prognostic model (PLANS), including five predictor variables of platelet count, lymphocyte count, age, neutrophil count, and sex.

This PLANS model showed excellent discriminative and calibration accuracy in predicting in-hospital mortality in COVID-19 patients.

The model would assist clinicians in better triaging patients and allocating healthcare resources to reduce COVID-19 fatality.

## Contributors

JL, YS, JJ and YY have full access to all the data in this study and takes full responsibility as a guarantor for the integrity of the data and the accuracy of the data analysis. JL, YC, SC, YS, JJ, and YY had the idea for and designed the study. SC, SW, DZ, HZ, YS and JJ collected the clinical data. JL, YC, SC, GQ and YY processed statistical data. JL, YC, YY drafted the manuscript. All authors interpreted the data and revised the manuscript critically. The corresponding author attests that all listed authors meet authorship criteria and that no others meeting the criteria have been omitted.

## Ethical approval

The study was approved by Jinyintan Hospital Ethics Committee (KY-2020-06.01), Union Hospital Ethics Committee (2020-0039), and the Institutional Review Board of Tongji Hospital, China (No. TJ-C20200140). Written informed consent was waived by these three Ethics Committees due to the emergence of this infectious diseases.

## Data sharing

No additional data available.

## Transparency

The manuscript is an honest, accurate, and transparent account of the study being reported; that no important aspects of the study have been omitted; and that any discrepancies from the study as planned have been explained.

## Copyright statement

The corresponding author of this article has theright to grant on behalf of all authors and does grant on behalf of all authors, an exclusive licence on a worldwide basis to the BMJ Publishing Group Ltd to permit this article (if accepted) to be published in BMJ editions and any other BMJPGL products and sublicences such use and exploit all subsidiary rights, as set out in our licence

## Supplementary

TRIPOD Checklist

Appendix S1. Implementation and estimates of Fine-Gray model

Appendix S2. Internal validation by bootstrap

Figure S1. Survival curves for derivation and validation cohort (One patient in the validation cohort was excluded due to missing LOS).

Table S1. “Baseline” survival probability (Wuhan, China)

Table S2. Thresholds and corresponding proportion and death toll included in each risk group

Table S3. Basic characteristics used in entropy balancing in Derivation cohort, New York cohort and Lombardy cohort

Table S4. “Baseline” survival probability (New York, USA)

Table S5. “Baseline” survival probability (Lombardy, Italy)

Table S6. Methodology quality assessment based on PROBAST risk of bias assessment tool

